# Profiling the human luminal small intestinal microbiome using a novel ingestible medical device

**DOI:** 10.1101/2025.04.18.25326056

**Authors:** Alexandre Tronel, Morgane Roger-Margueritat, Caroline Plazy, Salomé Biennier, Anthony Craspay, Ipsita Mohanty, Stéphanie Cools Portier, Manolo Laiola, Guus Roeselers, Nicolas Mathieu, Marianne Hupe, Pieter C. Dorrestein, Jean-Pierre Alcaraz, Donald Martin, Philippe Cinquin, Anne-Sophie Silvent, Joris Giai, Marion Proust, Thomas Soranzo, Elena Buelow, Audrey Le Gouellec

**Author notes:** Corresponding authors and co-last.

## Abstract

The invasive nature of sample collection for studying the small intestinal (SI) microbiome often results in its poor characterization. This study evaluated a novel ingestible medical device (MD) for SI luminal sample collection. A monocentric interventional trial (NCT05477069) was conducted on 15 healthy subjects. Metagenomics, metabolomics and culturomics assessed the MD’s effectiveness in characterizing the healthy SI microbiome and identifying potential biomarkers. The SI microbiota differed significantly from the fecal microbiota, displaying high inter-individual variability, lower species richness, and reduced alpha diversity. A combined untargeted and semi-targeted LC-MS/MS metabolomics approach identified a distinct SI metabolic footprint, with bile acids and amino acids being the most abundant classes of metabolites. Host and host/microbe-derived bile acids were particularly abundant in SI samples. The application of a fast culturomics approach to two SI samples enabled species-level characterization, resulting in the identification of 90 bacterial species, including five potential novel species. The present study demonstrates the efficacy of our novel sampling MD in enabling comprehensive SI microbiome analysis through an integrative multi-omics approach, allowing the identification of distinct microbiome signatures between SI and fecal samples.

## INTRODUCTION

The gut microbiota is defined as a complex microbial ecosystem comprising bacteria, fungi, viruses, protists and archaea. This ecosystem plays a direct role in four major functions that contribute to gut homeostasis and, consequently, to the health of the human host. The functions include metabolism^1^, regulation of the immune system^2^, endocrine functions^3,4^ and the regulation of organ physiology (e.g. gut-brain axis^5^, gut-liver axis^6^). Metabolites, which originate from gut microbes, the host or the diet, serve as the primary mediators of interactions between microbes and between the host and its microbes. The collective of all microorganisms, genetic material and biological compounds they contain and produce is referred to as the microbiome^7^. Environmental factors of the gastrointestinal tract (GIT), including oxygen concentration^8^, pH^9^, mucus^10^ gradients, and host secretions^11^ (e.g., pancreatic juice, bile, saliva and gastric juice), significantly influence the composition of gut microbiota. For instance, oxygen levels exhibit significant fluctuations along the GIT, reaching a minimum in the distal GIT and thereby establishing an anoxic environment. These fluctuations result in substantial variations in oxygen concentration between the small and large intestine^12^. Transit time and substrate availability within the GIT have been shown to significantly influence the composition of the gut microbiota^13^. Notably, distinct differences are observed between the small intestine (SI) and the colon. For example, the transit time through the SI typically ranges from two to five hours, resulting in a relatively dynamic environment compared to the slower and more stable conditions of the colon^14–16^. A variety of host secretions are secreted in the duodenum, the upper part of the SI, to allow degradation and absorption of food by pancreatic juice or bile. Bile acids (BAs) are metabolites that are produced by the host’s hepatocytes and secreted in the bile into the duodenum. They facilitate solubility and absorption of hydrophobic compounds and lipid emulsification^17,18^. BAs have been demonstrated to modify the gut microbiota by inducing damage to the bacterial membrane due to their surfactant properties and can cause DNA damage and oxidative stress on cells^19^. On the other hand, the gut microbiota plays a pivotal role in the biotransformation of BAs, for instance, through the deconjugation transformation^20^. Furthermore, there is evidence that the SI microbiota exhibits higher expression of genes involved in the transportation of simple carbohydrates, central metabolism, or biotin production when compared to the fecal microbiota ^13^.

Bacterial density and diversity increase progressively along the human GIT, ranging from 10^3^ CFU/mL in the upper intestinal tract to 10^11^ CFU/mL in the distal part of the gut^21^. The fecal microbiota has been the subject of extensive research and is predominantly composed of four main phyla: *Bacillota, Bacteroidota, Actinomycetota*, and *Pseudomonadota*^5^. However, it is important to note that the fecal microbiome is only partially representative of the proximal parts of the intestine. The SI microbiota is not well characterized at present, due to its difficult accessibility and the limited number of clinical studies that have investigated it^22,23^. The majority of these studies have collected samples that were derived from patients either during a surgical procedure^24^, endoscopy^25,26^, ileostomy^13^, or following an intestinal resection, or via a catheter aspiration^27^. Therefore, the development of medical devices (MD) capable of non-invasively collecting SI content is imperative to study the SI microbiome^28^. To date, some clinical investigations have evaluated the performance of such devices^29–32^ and demonstrated that the SI microbiome is less diverse than the fecal microbiome and dominated by the genera *Lactobacillus, Clostridium, Streptococcus, Staphylococcus, Veillonella* and *Bacteroides*^22–24,33–36^. However, knowledge about the SI metabolome remains limited, with most samples having been analyzed by non-targeted or targeted metabolomics on short chain fatty acids (SCFA) and BAs^29,37,37,38^. Culture-based analysis of SI content, which could eventually allow strain level characterization and have implications for the development of therapeutic probiotics and prebiotics, has been limited to cell viability counting and has included only the isolation of bacteria under aerobic conditions^29,35,39^. To the best of our knowledge, a culturomic approach that isolates and analyses bacteria from the SI under anaerobic and aerobic conditions using different media and culture conditions, has not yet been employed.

In this study, a first in man clinical investigation was conducted (detailed protocol described elsewhere^30^) to evaluate the safety and performance of a novel sampling MD that collects intestinal content from the SI in a non-invasive manner. The technology utilizes a pH-dependent enteric capsule containing three sampling modules, enabling the collection of triplicate samples. The secondary objectives of the study were to perform multi-omics analysis to characterize the upper intestinal microbiota composition and metabolome with a focus on the identification of potential biomarkers specific for the SI. In this study, we initially demonstrated the feasibility of conducting multi-omic analyses (metagenomics, metabolomics and culturomics) on SI contents. By analyzing the corresponding feces, we demonstrated significant differences in microbial composition and metabolites between SI contents and fecal samples. The analysis of SI contents revealed the presence of specific metabolites, including glycocholic acid and taurocholic acid, which are host-derived BAs. Furthermore, culturomics was employed to characterize and archive 90 bacterial species including five previously uncharacterized species. This comprehensive approach underscores the efficacy of the MD for elucidating the intricate composition of the SI microbiome in healthy individuals and for the prospective identification of novel biological markers.

## MATERIALS AND METHODS

### Description of the sampling medical device

The sampling MD employed in this clinical investigation was developed by Pelican Health SAS, La Tronche, France (patent number: WO2019081539)^40^. The device is designed for single-use, is ingestible and is minimally invasive. It is intended for the collection of samples from the luminal small intestinal fluid. The device consists of a gastro-resistant and pH-dependent pill (size 00) for enteric use and three sampling modules with a final size of 8mm x 22mm (See Figure 1). Each sampling module is composed of silicone and contains a compressed super-absorbent polymer (comparable to a sponge) that functions as a reservoir. The size of a sampling module is 6.5mm x 7mm. Upon the introduction of fluid into the module, the compressed polymer undergoes expansion, resulting in the release of mechanical energy that activates the closing of valves. This process effectively isolates the sample from the external environment. The complete modules are then transferred through the large bowel and subsequently collected in the feces for further analysis.

**Figure 1:**
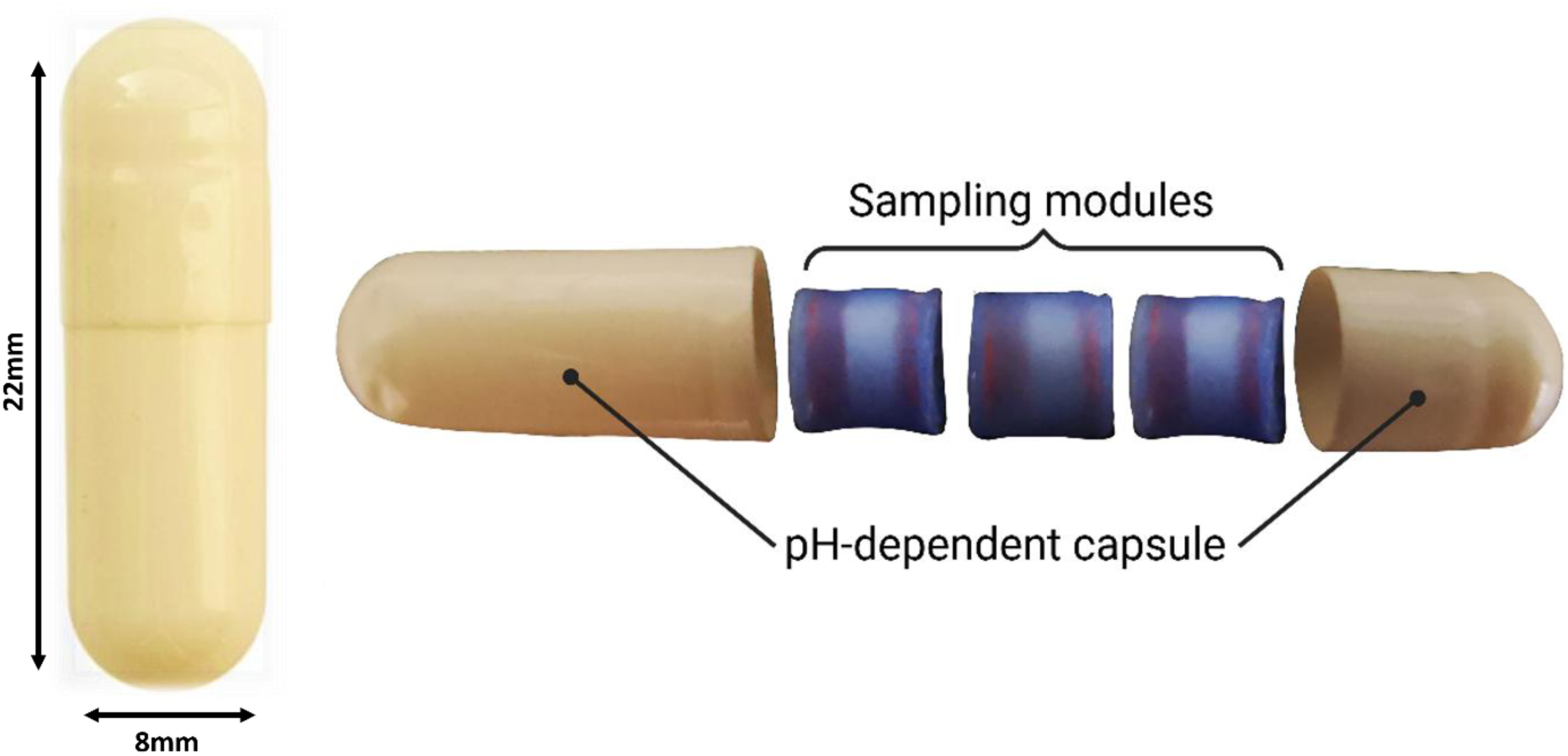
Photograph of the sampling MD evaluated in this clinical investigation.

### Participants and study design

The clinical investigation protocol has been previously described in more detail^30^. The main inclusion criteria for healthy volunteers were: subjects must (1) be aged between 18 and 65 years; (2) have a body mass index (BMI) greater than 20 and lower than 30 and (3) be affiliated with a social security system. A large number of exclusion criteria were implemented to ensure that the subjects had no gastro-intestinal issues or any co-morbidities^30^. In addition, candidates taking medication (antibiotics) or probiotics and related substances were excluded. The protocol was approved by the Personal Protection Committee (CPP) and the French National Agency for the Safety of Medicines and Health Products (ANSM).

A total of 15 healthy volunteers were included during the study and each subject swallowed one MD under the supervision of a gastroenterologist following a 10 hour fast. Upon reaching the jejunal/ileal part of the SI^41^, the pill released three sampling modules to collect luminal content. Fecal samples were collected in a designated collection device and stored in a cool box immediately after collection before being directly transferred to the Grenoble’s hospital. The global workflow for sample management is described in more detail in the published protocol^30^. No modules were detected in the feces of one volunteer, consequently only samples for 14 of the 15 volunteers were analyzed.

### Sample preparation for multi-omics analysis

Following collection, fecal samples were directly processed at the hospital. Initially, the feces were screened for the presence of the collection modules released from the MD. In the absence of a detected module, the subject’s fecal collection was continued. Conversely, if at least one module was detected in the collected fecal sample, the modules were recovered and separately processed to assess the volume sampled. The upper intestinal content was then retrieved by means of centrifugation. The bacterial pellets were subsequently utilized for metabarcoding sequencing and while the residual fluids were allocated for metabolomics analysis. Concurrently, fecal samples were also processed to perform the same analysis (metabarcoding and metabolomics respectively). Additionally, for only two subjects, a culturomics approach was performed on intestinal content. All samples were stored at −80°C until analysis, except for the culturomics samples.

### Determination of bacterial composition by metabarcoding sequencing

#### DNA extraction

For fecal samples, extraction was performed on 250 mg of sample. Conversely, DNA was extracted by an external service provider (ADM biopolis, Valencia, Spain) on the whole bacterial pellet retrieved from the SI content after centrifugation (15 min 15 000g). The protocol of the commercial PowerFecal Pro DNA Kit by Qiagen (Qiagen, Hilden, Germany) was used for DNA isolation. Briefly, a cell lysate was obtained by mechanical disruption (Fastprep, Thermo Fisher Scientific) and chemical treatment. The isolation of DNA and the purification of contaminants and inhibitors was performed using a silica column. For samples from intestinal content yielded very low concentration DNA, an extra step of purification was done following with QIAamp DNA Micro Kit (Qiagen, Hilden, Germany). Photometric (UV) evaluation was used to assess DNA quality and concentration using a Nanodrop apparatus (Thermo Fisher Scientific, Waltham, MA, USA).

### 16S rRNA sequencing analysis

The metabarcoding, sequencing and bioinformatic processing were carried out by an external service provider (ADM biopolis, Valencia, Spain). The DNA concentration used for amplification was 50 ng, following the 16S Metagenomic Sequencing Library Illumina 15044223 B protocol (Illumina). In summary, in the first amplification step, the region of interest has been amplified (V3-V4) with 16S rRNA gene universal primers^42^ and a universal linker sequence allowing amplicons to incorporate indexes and sequencing primers during the secondary PCR step. The quantification of 16S-based amplicon was performed by fluorimetry using the Quant-iT™ PicoGreen™ dsDNA Assay Kit (Thermo fisher Scientific, Waltham, MA, USA). In the second assay amplification indexes Nextera XT Index kit (Illumina) were included. The quantification of 16S-based libraries was performed by fluorimetry using the Quant-iT™ PicoGreen™ dsDNA Assay Kit (Thermo fisher Scientific, Waltham, MA, USA). Libraries were pooled (equimolar). The size and quantity of the pool were assessed on the Bioanalyzer 2100 (Agilent) and with the Library Quantification Kit for Illumina (Kapa Biosciences), respectively. PhiX Control library (v3) (Illumina) was combined with the amplicon library (expected at 20%) prior to sequencing on the MiSeq platform (Illumina), 300 cycles in paired reads configuration. Image analyses, base calling, and data quality assessment were performed on the MiSeq instrument. Raw sequences, forward and reverse, were merged in order to obtain the paired-end sequences using the BBMerge package of BBMap V.38 software with minimum 70 nt overlap on each end. The amplification primers from the sequences obtained in the sequencing step were trimmed to reduce bias in the annotation step, with Cutadapt v 1.8.1 and parameters by default. Once the primers were removed, sequences lower than 200 nt were removed from the analysis. After obtaining the clean complete sequences using Reformat module from BBMap v.38, a quality filter was applied to delete poor-quality sequences. Those bases in extreme positions that did not reach Q20 (99% well-incorporated base in the sequencing step) or a greater phred score were removed. Subsequently, sequences whose average quality did not surpass the Q20 threshold, as a mean quality of the whole sequence, were also deleted. The reads were processed using the DADA2^43^ denoise-single command. Error rates were learned from a set of subsampled reads using “learnErrors” and a sample inference algorithm was applied with the “dada” function. The chimeric amplicon sequence variants (ASVs) were removed using “removeChimeraDenovo”. Those clean ASVs were annotated against the NCBI 16S rRNA database version 2021 using Blastn version 2.2.29+^44^. The taxonomy of the ASVs that had been assigned with a lower percentage identity than 97%, was reassigned using NBAYES algorithm^45^. NBAYES classifier was trained on V3-V4 regions of 16S rRNA gene from SILVA v.138 database^46^.

### Determination of the metabolomes

#### Sample preparation for the untargeted and semi-targeted metabolomics

Two distinct samples types were prepared for Liquid Chromatography-High Resolution Mass Spectrometry tandem (LC-HRMS/MS) acquisition: the SI content sampled with the MD and the fecal samples.

For SI content, metabolites were extracted by the addition of cold methanol (MeOH) (1:4 (v/v)) spiked with deuterated leucine (d-leucine) for protein precipitation and then vortexed for 20s. For fecal samples, first the water content was determined by drying a part of the sample in an oven. Separately, a total of 150 mg of wet feces was used for metabolomic analyses. The water content was corrected by adding water to samples (correction based on the higher water content). Samples were then homogenized by vortex and sonication for 10 min (on ice). MeOH spiked with d-leucine was added (1:4 (w/v)) for metabolites extraction and protein precipitation. Fecal and intestinal samples were then incubated on ice for 30 min and centrifuged 15 min, 15 000 g, +4°C. After separation of supernatants and pellets, supernatants were evaporated under nitrogen flow at room temperature. The metabolite dry pellets were resuspended in liquid chromatographic solvent (80% water, 20% MeOH, 1% acetonitrile (ACN), 0,1% formic acid (FA) and internal standards (deuterated tryptophan and deuterated phenylalanine)). Resuspension volumes were different for each SI sample to normalize the concentration of intestinal fluid injected during the analysis.

#### Combined untargeted and semi-targeted metabolomics approach

A combined untargeted and semi-targeted metabolomics approach was applied using an Ultra-high-performance liquid chromatography (UHPLC) (Vanquish Flex, Thermo Fisher Scientific, Waltham, MA, USA) coupled with a Q Exactive Plus Orbitrap HRMS/MS (Thermo Fisher Scientific, Waltham, MA, USA)^47^. Thirty-seven metabolites of interest that have been previously selected^47^ (listed in Supplementary Table 1), have been relatively quantified with the semi-targeted method.

The chromatographic separation was carried out on a Luna Omega polar C18 (2.1mm × 150 mm, 1.6 μm, Phenomenex, Torrance, CA, USA) at 30 °C with a flow elution rate of 400 μL/min. The temperature of the autosampler compartment was set at +4°C, and the injection volume was 5 μL. For the chromatographic settings, the mobile phases consisted of A (water + 0.1% FA) and B (ACN + 0.1% FA). Elution started with an isocratic step of 2 min at 1% mobile phase B, followed by a linear gradient from 1% to 100% mobile phase B for the next 12 min. The next 6 min were with 100% mobile phase B before returning to 1% B for 5 min corresponding to the column equilibration. The entire chromatographic acquisition lasted 25 min. The mass spectrometer was fitted with an electrospray source (ESI) operating in positive and negative ionization modes. The scan range was from *m/z* 85.0 to 1275.0 with a resolution of 70 000 at a ratio *m/z =* 200. MS2 fragmentation was performed at three collision energies (CEs: 10, 35, 55 eVs) on the top 10 of the most abundant parent ions during full scan. Quality controls (QC) were used to ensure the quality of the LC-HRMS/MS acquisition. Deuterated compounds were used for controlling extraction process (d-leucine) and injection (d-tryptophan and d-phenylalanine). Pooled QC assembled from all biological samples ensured the stability of peak detection and intensity. They were injected every 10 injections. Briefly, for the semi-targeted metabolomics, an external calibration curve with the pooled 37 metabolites of interest was injected at different dilution factor, allowing to estimate the concentrations of each metabolite in the biological samples^47^.

Data were then analyzed for untargeted metabolomics MZmine 3.9.0^48^ as described elsewhere^49^. We evaluated the threshold’s noise on our instrument in full scan at an intensity of 1E6 and 1E3 in MS2. After data processing on MZmine3, data was filtrated based on a coefficient of variation for each feature on the QC pool lower than 30% and the feature must be present in 10% of the total of biological samples. Feature annotation was achieved using Feature Based Molecular Networking (FBMN)^50^ on Global Natural Products Social Networking 2 (GNPS2).

The semi-targeted analysis was performed using the Thermo Fisher Scientific software TraceFinder 4.1 General Quan and has been previously described^47^. Briefly, after the construction of the compound database containing the 37 metabolites of interest, samples were analyzed by the software to find the metabolites based on full scan spectra, retention time and MS2 spectra. Data containing peak areas for each sample were then exported in .csv file for deeper analysis. Precisely, sample losses due to the metabolite extraction method were assessed by measuring the recovery percentage by dividing the detected d-leucine in each sample by the expected d-leucine quantity. Then the following formula is applied to obtain metabolite quantity in arbitrary unit per mg for feces or µL for intestinal liquid: peak area / (recovery*mg for feces or µL injected). These data were plotted to obtain boxplots for feces or intestinal liquid for each metabolite with Matplotlib v. 3.5.1^51^ in Python v. 3.9.5.

To detect as many BAs as possible, we reprocessed the untargeted metabolomics data by reducing filtration. BA annotations and relative quantification using peak area abundances extracted using MZmine 4.1.0^48^ were achieved using FBMN^50^ on GNPS2 and an expanded set of bile acid libraries^52,53^.

#### Sample preparation for the targeted metabolomics on bile acids

A fraction of the samples from feces and SI content were mixed with MeOH (4:1) and homogenized with a bead beater for 4 min. Subsequently, they were centrifuged at 16 000 g at 4°C for 10 min. The supernatant was then transferred to Phree filters and centrifuged at 15 000 g at 4°C for 5 min. The filtrates were stored at -20 °C until analysis.

#### Targeted metabolomics analysis on bile acids

The analysis of BAs was performed by an external provider (Clinical Microbiomics, Vedbæk, Denmark) and was carried out in a randomized order using a UHPLC system (Vanquish, ThermoFisher Scientific, Waltham, MA, USA) coupled with a high-resolution mass spectrometer Q Exactive™ HF Hybrid Quadrupole-Orbitrap, (Thermo Fisher Scientific, Waltham, MA, USA). An electrospray ionization interface was used as an ion source in negative mode. The chromatographic separation of bile acids was carried out on a Waters Acquity HSS T3 1.8 μm 2.1 x 150 mm (Waters). The column was thermostated at 30°C. The mobile phases consisted of (A) ammonium acetate 10 mM, and (B) MeOH/ACN (1:1, v/v). Bile acids were eluted by increasing B in A from 45 to 100 % for 16 min. Flow rate was 0.3 ml/min. To ensure high-quality sample preparation, a quality control sample (QC sample) was prepared by pooling small equal aliquots from each sample, to create a representative average of the entire set. This sample was treated and analyzed at regular intervals throughout the sequence. Possible matrix effects on compounds for quantification, were tested by spiking aliquots of the QC sample at a minimum of two levels. A QC sample was analyzed in MS/MS mode for the identification of compounds. Peak areas were extracted using Skyline 23.1 (MacCoss Lab Software). Identification of compounds was based on accurate mass and retention time of authentic standards. The concentrations of 50 BAs have been determined. The BA list is available in Supplementary Table 2.

### Determination of microbial composition by a fast culturomics approach

#### Media preparation

Different media were used for fast culturomics. The first one is Yeast Casitone Fatty Acids (YCFA) medium (Deutsche Sammlung von Mikroorganismen und Zellkulturen, DSMZ_medium1611). Liquid YCFA was degassed with argon gas for 15 min and autoclaved before filling into Hungate tubes (10 mL) and supplemented with 2mL of anaerobic sterile rumen juice and 2 mL of anaerobic sterile defibrinated sheep blood (SARL Atlantis, Voulmentin, France). For solid YCFA, the medium was enriched with 5% of sterile rumen juice and 5% of defibrinated sheep blood. Sterile rumen juice was prepared following the procedure described by Diakite *et al.*^54^. The other solid media were sheep blood agar plate (COS) (BioMérieux, Marcy l’Etoile, France) and Sabouraud/Chloramphenicol agar plate (BioMérieux, Marcy l’Etoile, France). In addition, two other liquid media were used: anaerobic culture vial medium and aerobic culture vial medium from BD BACTEC^TM^ (Becton Dickinson, Franklin Lakes, NJ, USA). These media were supplemented with 2 mL of sterile rumen juice and 2 mL of defibrinated sheep blood.

#### Fast culturomics protocol

Culturomics was performed on two intestinal samples just after the module’s collection in feces. To be used for culturomics, intestinal samples must meet criteria such as have a sufficient collected volume >50µL and no fecal contamination (dark spot on the polymer).

The fast culturomics protocol (Figure 2) was adapted from the fast culturomics method developed by Naud *et al.*^55^. In summary, the small intestinal content was placed immediately after collection in an anaerobic bag and culturomics process started within one hour. Following gentle centrifugation and phosphate buffer saline (PBS) washes (two cycles) (at the conclusion of which the sample was 10-fold diluted), the intestinal suspension was placed in an anaerobic chamber (Whitley A35 anaerobic station, Don Whitley scientific, Bingley, England). The gas alimenting the anaerobic chamber was composed of 90% nitrogen, 5% hydrogen and 5% carbon dioxide. The experimental design incorporated a range of culture conditions. First, 100 µL of the 10 times diluted sample are used for a series of dilutions (from 10^-1^ to 10^-5^) in PBS and 50 µL of each dilution are plated on YFCA modified, COS and Sabouraud/Chloramphenicol agar plates. These direct cultures were then subjected to incubation for a period of 48 hours at a temperature of 37°C, within either an anaerobic or an aerobic atmosphere. Concurrently, pre-cultures (liquid enrichment) have been performed in YCFA-modified liquid medium and anaerobic/aerobic culture vial media. These pre-cultures were then subjected to incubation at 37°C within either anaerobic or aerobic atmospheres without agitation. Sub-culturing was performed after three hours, six hours, nine hours, 24 hours, 72 hours, seven days, and 10 days of incubation. To this end, a dilution series ranging from 10^-2^ to 10^-8^ was established using 100 µL of the pre-culture. These dilutions (50 µL) were subsequently plated onto YCFA agar plates and COS agar plates, with the former being used for the YCFA pre-culture and the latter for the anaerobic/aerobic culture vial media. These plates were then subjected to an incubation period of 48 hours at a temperature of 37°C within either an anaerobic or an aerobic atmosphere.

**Figure 2:**
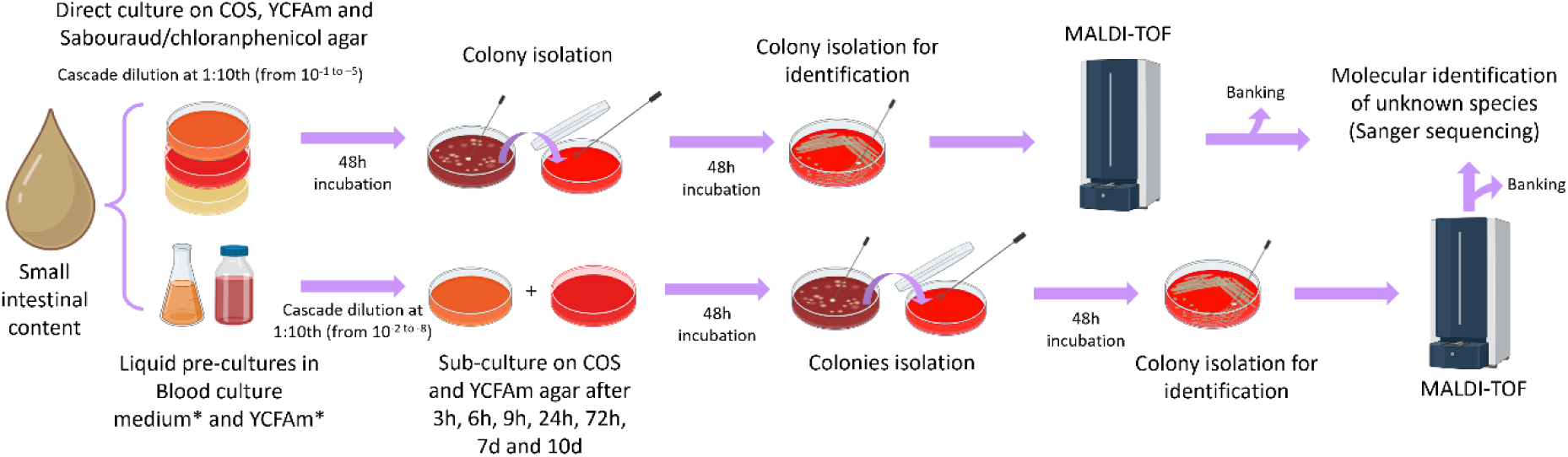
Culturomics workflow in anaerobic and aerobic conditions performed on small intestinal contents (the figure was created in BioRender).

#### Microbial identification

The isolation of microbial colonies was conducted following a 48-hour incubation period, contingent on their distinct morphological characteristics, including size and color. Isolates were then streaked again on the same agar plate from which they were initially isolated (YCFA or COS). Following an additional 48 hours of incubation, identification of the isolates was conducted through the use of mass spectrometry Matrix Assisted Laser Desorption-Time of Flight (MALDI-TOF). The MALDI-TOF MS Biotyper (Bruker Daltonik, Bremen, Germany) was utilized for this purpose. The obtained spectra were then compared to the MBT IVD Library Revision J (2022), which contains spectra from 4,194 microbial species. Colony identifications were considered valid if the identification score was ≥1.8. In instances where the identification score was less than 1.8, a secondary MALDI-TOF analysis was conducted to ascertain the accuracy of the initial identification. If the score fell below 1.8 on the second attempt, the isolate was designated as "unknown."

The identification of unknown isolates necessitated the implementation of full 16S rRNA gene sequencing. Genomic DNA was extracted using the PowerFecal Pro DNA Kit by Qiagen (Qiagen, Hilden, Germany). Full 16S rRNA gene DNA was amplified by PCR using the following primers: F: 5’-AGRGTTYGATYMTGGCTCAG and R: 5’-CGGYTACCTTGTTACGACTT and the KAPA2G Robust HotStart ReadyMix (Kapa Biosystems, Wilmington, MA, USA). The PCR conditions were as follows: initial denaturation at 95°C for 3 minutes, denaturation at 95°C for 15 seconds, annealing at 60°C for 15 seconds, extension at 72°C for 60 seconds and final extension at 72°C for 2 minutes. A total of 30 cycles were completed. Subsequently to this, the amplicons were then purified using the magnetic beads purification kit AMPure, and the DNA was quantified with a Nanodrop (Thermo Fisher Scientific, Waltham, MA, USA). Subsequently, Sanger sequencing was performed using the same primers as the PCR. The QV30 sequences were then subjected to analysis with the aid of the Benchling software, wherein the forward and reverse sequences were aligned to construct the consensus sequence (alignment program MAFFT^56^). The consensus sequences were then analyzed using EZbiocloud^57^. The definition of a new species was based on a 16S rRNA gene sequence similarity of less than 98.65% with the closest relatives^58^.

Isolates were banked in YCFA or anaerobic culture vial media and 20% glycerol and stored at -80°C.

### Statistical analysis

All statistical analyses were performed using R software with *vegan*^59^ package for diversity analysis and *phyloseq*^60^ for microbiome data handling. Alpha diversity was evaluated as species richness, defined as the number of different ASVs, and species diversity via the Shannon index, which integrates richness and evenness. Statistical comparisons were conducted using a paired Wilcoxon rank-sum test, with a p-value threshold set at < 0.05. The beta diversity (differences in microbial composition) between SI content and fecal samples was evaluated using Bray-Curtis dissimilarity matrix and visualized via Principal Coordinate Analysis (PCoA). The statistical significance of the observed differences in the PCoA was evaluated using PERMANOVA (Permutational Multivariate Analysis of Variance) with 999 permutations, and a p-value threshold of < 0.05. To assess the variability and interindividual variation within groups PERMDISP (Permutational Analysis of Multivariate Dispersions) was used to evaluate the homogeneity of group dispersions. This was achieved by calculating the distance of each sample to the group centroid and comparing these distances between groups.

In order to ascertain the bacterial genera that potentially serve as biomarkers of biological matrices, the fold change (FC) (SI+1/feces+1) was determined for each bacterial genus from the normalized relative abundance data. The analysis was conducted on genera present in a minimum of 20% of the intestinal content or fecal samples. The Wilcoxon statistical test was employed to compare the mean relative abundances between the two groups. P-values were adjusted by using the Benjamini-Hochberg procedure with the False Discovery Rate (FDR).

For the untargeted metabolomic approach, MetaboAnalyst 6.0^61^ was utilized. Normalization of samples was achieved through median normalization, and data were log-transformed. To ensure data integrity and enhance interpretability, Pareto scaling was employed for data scaling. Volcano plots were generated using MetaboAnalyst 6.0, and the top 30 features were annotated on the plot. The selected FC (small intestine/feces) threshold was 2, and the p-value threshold was 0.05.

For semi-targeted and BAs targeted metabolomics, statistical analysis was applied to assess significant differences between feces and intestinal content thanks to a non-parametric Wilcoxon test. In this case, p-values were corrected by using the Benjamini-Hochberg procedure with the False Discovery Rate (FDR) obtained.

## RESULTS

Utilizing our MD, we obtained samples of SI content and fecal samples from 14 healthy volunteers and compared their microbiota and metabolic profiles by metagenomic barcoding, culturomics and metabolomics.

### The SI microbiota differs from the fecal microbiome

The bacterial composition of the samples collected with the MD and their corresponding fecal samples was first studied by 16S rRNA gene sequencing. A significant difference in species richness (p-value < 0.0001) was identified between the two sample types, with fecal samples exhibiting a higher richness (Figure 3a). Microbial diversity (Shannon index) differed between sample types. Specifically, the SI microbial composition exhibited significantly lower diversity (p<0.0001) compared to that of the fecal samples (Figure 3b). Principal coordinate analysis (PCoA) on ASV level (Bray-Curtis distances) indicated significant differences in microbial composition between SI content and feces (p<0.001) (Figure 3c). The PERMDISP test revealed a greater dispersion within the SI content group compared to the fecal samples. This finding indicates that the SI content group exhibited higher variability, suggesting the presence of greater inter-individual variation in the upper intestinal tract (Supplementary Figure 1).

**Figure 3:**
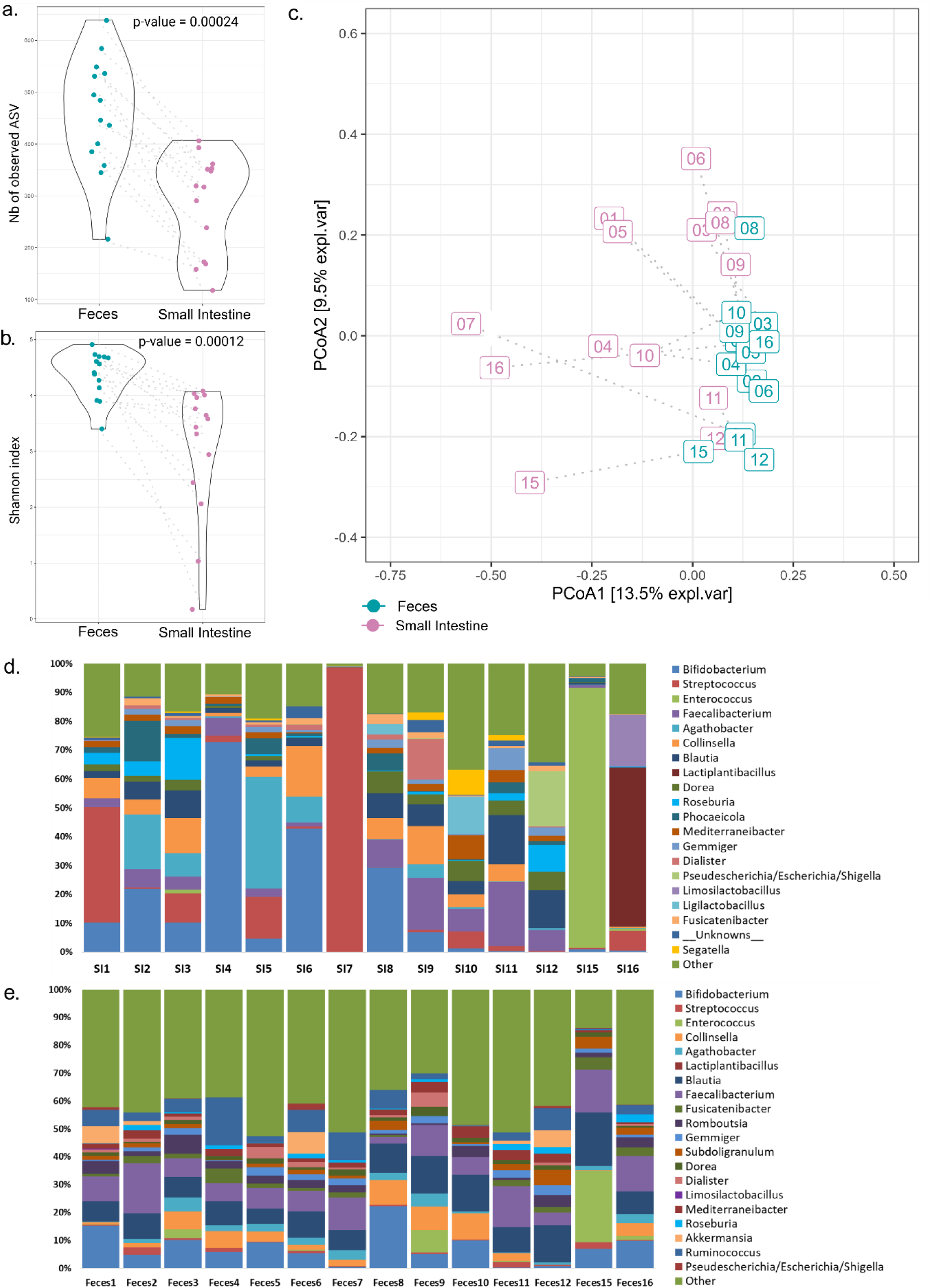
Microbial composition of the small intestinal contents (n=14) and fecal samples (n=14). (a) Bacterial richness and (b) bacterial diversity in small intestinal contents and fecal samples. The samples (SI content vs feces) for each subject are connected by dotted lines. (c) Microbial community β-diversity at ASV level, which was demonstrated using principal coordinates analysis (PCoA) of the Bray-Curtis distance matrix. The samples (SI content vs feces) for each subject are connected by dotted lines. (d) Relative abundance of the 20 most predominant bacterial genera in small intestinal contents. Remaining genera are summarized as “Other”. Each column represents one subject. (e) Relative abundance of the 20 most abundant bacterial genera in fecal samples. Other genera are summarized as “Other”. Each column represents one subject.

*Bacillota* was the most abundant phylum (70%) detected for the two groups of samples (SI and feces respectively). Differences were observed for the mean relative abundance of *Bacteroidota* and *Actinomycetota*, with a higher level in feces (14% vs 5,6%) and a higher level in SI content (21,3% vs 12,9%), respectively. At the genus level, a higher abundance of bacteria from the genera *Streptococcus* and *Bifidobacterium* was observed in the SI contents compared to the fecal samples (Figure 3d and 3e). A notable degree of inter-individual variation in SI contents was evident. This observation was particularly pronounced for individuals exhibiting a remarkably elevated relative abundance of *Streptococcus* (SI7), *Bifidobacterium* (SI4), or *Enterococcus* (SI15) in their respective samples (Figure 3d). Overall, fecal samples showed less inter-individual variation except for one subject (S15) with a high level of *Enterococcus* (relative abundance of 90% in SI content and 25% in feces) (Figure 3e). We calculated the relative abundance fold change (FC) for each genus to identify the most discriminating genera for the respective sample types (on average). For SI content, bacteria from the genera *Schaalia* (FC=1.24, q-value<0.05), *Gemella* (FC=1.19, q-value<0.05) and *Granulicatella* (FC=1.11, q-value<0.05) were the most discriminant genera (Supplementary Table 3). On the other hand, *Ruminococcus* (FC=0.22, q-value<0.01)*, Bacteroides* (FC=0.34, q-value<0.01)*, Romboutsia* (FC=0.37, q-value<0.01) and *Akkermansia* (FC=0.44, q-value<0.05) were the most discriminant genera for feces (Supplementary Table 3). These genera may serve as potential indicators of the SI and feces microbiota respectively, in healthy individuals.

### Combination of untargeted and semi-targeted metabolomics reveals significant differences between the SI and the fecal metabolome

We also performed combined untargeted and semi-targeted metabolomics to extensively characterize the SI content versus its corresponding fecal metabolome^47^. In the positive ionization mode for untargeted metabolomics, we obtained 5289 features after processing with MZmine3. After filtering the data, 574 features remained, representing 10.8% of the original features. Principal component analysis (PCA) on the filtered data revealed a clear separation between SI contents and feces (p<0.001) (Figure 4a). Thus, the metabolomic profiles of the SI contents differed substantially from those of the fecal samples (R-squared: 0.63595).

**Figure 4:**
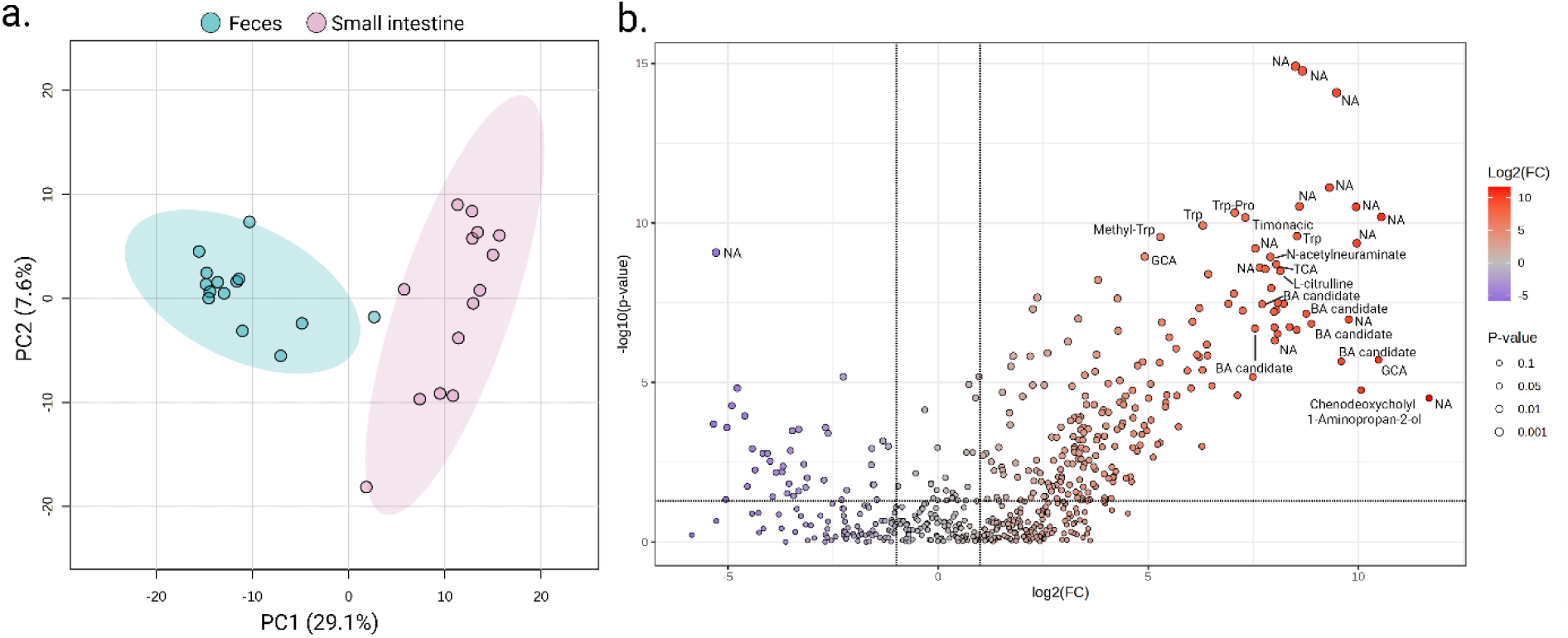
Comparison of metabolite abundance between the SI and fecal samples revealed significant differences across a wide range of compounds. (a) Principal Component Analysis (PCA) of the filtered features (*n*=574 metabolites) obtained in positive ionization mode. To assess statistical differences between the metabolomics profiles, PERMANOVA based on 999 Monte Carlo permutations was performed (F-value: 45.419; R-squared: 0.63595; p-value permutations: 0.001). (b) Volcano plot of the metabolites showing significant differences in the abondance of metabolites between the SI and the feces groups using a fold change (FC) threshold of 2 and a *t*-test threshold of 0.05. The log transformed FC and P-values are represented on x- and y-axes.

To identify abundance differences in metabolites between the two sample types, we performed supervised statistical analyses. The volcano plot based on the 574 features shows 206 features which were more abundant in SI contents (p<0.05) and 48 features more abundant in feces (p<0.05). From the 574 features, 189 could be annotated to the level 2 of annotation, meaning that the MS2 spectra matched to spectrum libraries^62^ (i.e, 33%). A total of 45% of the features were annotated among those with a p-value ≤ 0.05 and an FC ≥ 2.115. The largest chemical annotated subclasses were amino acids/peptides and BAs (Supplementary Table 4). A differential analysis of the metabolites revealed that certain metabolites, including BAs (glycocholic acid [GCA] and taurocholic acid [TCA]), as well as some candidate BAs, as well as tryptophan, methyl-tryptophan, and citrulline, exhibited higher concentrations in the SI content compared to feces (Figure 4b).

For semi-targeted metabolomics, 37 metabolites were targeted in both, the SI content and fecal samples. A total of 15 metabolites were found to be significantly more abundant in SI content compared to feces, particularly those derived from amino acids (tryptophan, phenylalanine, and leucine/isoleucine) and from the BA group (Figure 5). Conversely, seven metabolites were significantly more abundant in feces compared to SI content (e.g. biotin (p<0.001), nicotinic acid (p<0.001) and serotonin (p<0.01)). For instance, the mean concentration of tryptophan in SI content was estimated to be 1.15 x 10¹⁰ unit/µL and 7.16 x 10⁸ unit/mg in feces, and the mean concentration of nicotinic acid (vitamin B3) was estimated to be 1.39 x 10⁸ unit/µL and 2.78 x 10⁹ unit/mg in feces (Figure 5). These findings suggest that these concentrations may be indicative of their average concentration in healthy individuals.

**Figure 5:**
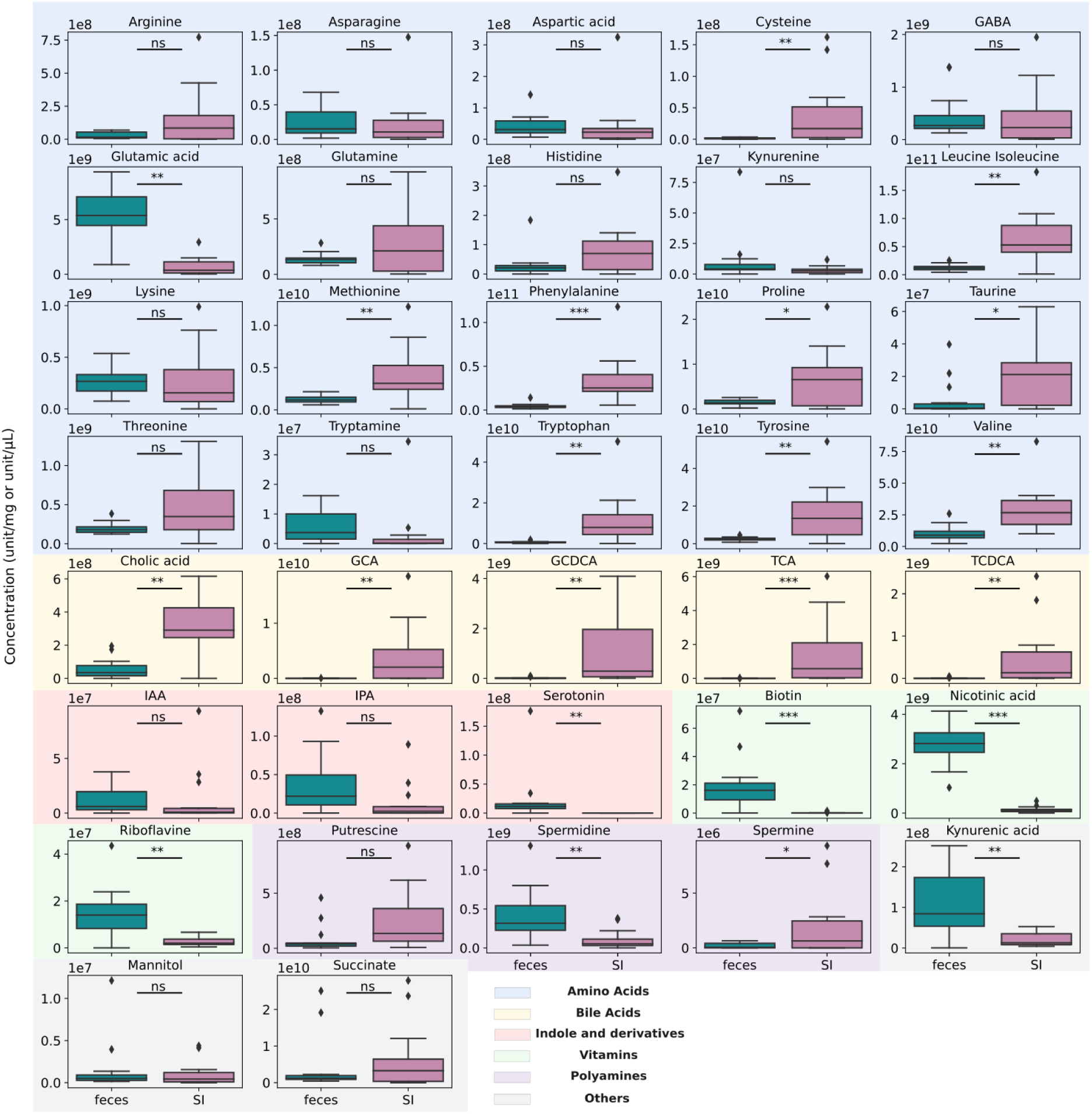
Relative quantification by semi-targeted metabolomics of the metabolites of interest in small intestinal contents and fecal samples (in unit/mg for fecal and unit/µL for small intestinal content). GCA: Glycocholic acid; GCDCA: Glycochenodeoxycholic acid; TCA: Taurocholic acid; TCDCA: Taurochenodeoxycholic acid; IAA: Indole-3-acetic acid; IPA: Indole-3-propoinic acid. *Indicates significant differences (p<0.05) between groups, **(p-0.001) and ***(p<0.0001).

### The SI microbiome: a significant reservoir for BAs

The application of targeted metabolomics on BAs revealed no statistically significant disparities between the SI content and the corresponding fecal samples, with respect to all targeted BAs (50 BAs). However, a significant difference was identified between the SI content and feces when BAs were categorized based on their source (host vs. microbe-derived BAs, respectively) (Figure 6a). The analysis revealed that microbe-derived BAs exhibited a marked increase in concentration in fecal samples (p<0.0001), while host-derived BAs demonstrated a significantly higher concentration in the SI content (p<0.01). Furthermore, BAs that can derive from both the host and microbes (host/microbe-derived BAs) were found to be more abundant in the SI content (p<0.058) (Figure 6a). The absolute concentrations for 50 BAs were subsequently determined (Figure 6b and Supplementary Figure 2). The mean concentrations of primary deconjugated BAs, cholic acid (CA) and chenodeoxycholic acid (CDCA), were evaluated to be 27.5 µM and 13.9 µM in the SI contents and 3.5 µM and 13.4 µM in feces, respectively. For glycine and taurine-conjugated BAs, the mean concentrations of GCA, glycochenodeoxycholic acid (GCDCA), TCA, and taurochenodeoxycholic acid (TCDCA) were found to be significantly higher in the SI contents compared to feces (23.7 µM in SI contents versus 0.01 µM in feces, 3.9 µM in SI contents versus 0.2 µM in feces, 7.4 µM in SI contents versus 0.01 µM in feces, and 1.6 µM in SI contents versus 0.01 µM in feces, respectively (Figure 6b). Conversely, secondary BAs were more abundant in feces. For example, deoxycholic acid (DCA) and lithocholic acid (LCA) mean concentrations were measured at 3.44 µM in SI contents versus 17.3 µM in feces and 0.2 µM in SI contents versus 30.5 µM in feces, respectively. The present study offers an initial overview of BA concentrations in healthy subjects (Supplementary Table 5). We determined that the median host-derived BA concentration was 21.56 µM in SI samples and 0.01 µM in feces. The global BA profile of each sample was determined using an untargeted metabolomics approach. Distinct profiles were identified between the two sample types, with a higher abundance of glycine and taurine conjugated BAs in SI content (Figure 6c). Conversely, fecal samples exhibited a higher abundance of Keto-BAs and candidate BAs derived from a novel BA library^52^. The analysis revealed interindividual variations within SI contents and feces groups, with five subjects exhibiting remarkably low abundances of glycine and taurine conjugated BAs in their SI contents. Utilizing the untargeted data, we observed a robust cluster of unconjugated BAs that exhibited higher levels in feces compared to the conjugated BAs present in the SI (Figure 6d). However, there were a few exceptions, including the histidine and alanine conjugated BAs, which appeared to be elevated in feces. Furthermore, sulphated deoxycholic acid levels were found to be elevated in the SI. We performed a molecular network analysis on BAs using FBMN on GNPS2 using the untargeted data (Supplementary Figure 3). MS2 spectral matches to 68 BAs were obtained by matching to the BILELIB19 library, which increased to 556 annotations by matching to the recently generated candidate bile acid library^52^. Matches to amino acid and other amine conjugated BAs were also recovered, with varying distribution between SI contents and feces.

**Figure 6:**
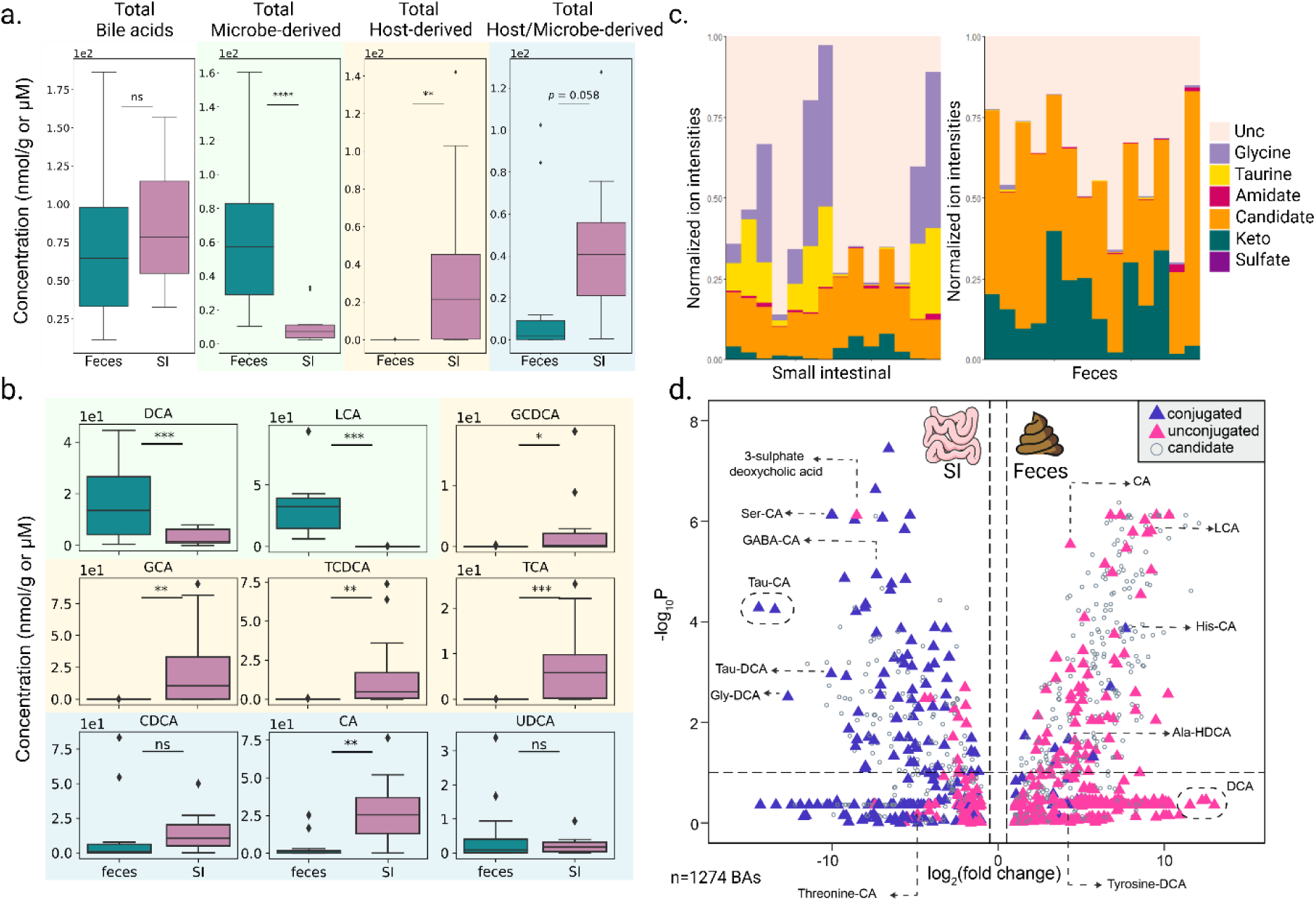
Determination of the Bile Acids (BAs) profile in SI contents (n=14) compared to their corresponding fecal samples (n=14). (a) Boxplots of the overall concentration of BAs in SI (n=14) and fecal (=n=14) samples. Concentration of total Microbe-derived BAs and total Host-derived BAs were depicted and compared separately as well as total Host/microbe-derived BAs (nmol/mg for fecal samples and µM for SI contents). (b) Boxplots of the most common BAs quantified in SI content or fecal sample. Microbe-derived BAs colored in green, Host-derived BAs colored in yellow and Host/Microbe-derived BAs colored in blue. DCA: Deoxycholic acid; LCA: Lithocholic acid; GCDCA: Glycochenodeoxycholic acid; GCA: Glycocholic acid; TCDCA: Taurochenodeoxycholic acid; TCA: Taurocholic acid; CDCA: Chenodeoxycholic acid; CA: Cholic acid and UDCA: Ursodeoxycholic acid. (c) Global profile of bile acids in SI contents and fecal samples. Each column represents a given sample from one subject. BAs have been grouped according to their chemical family, Unc: unconjugated BA. (d) Volcano plot demonstrating the log2 fold changes in the peak area abundances of the 14 subjects between their SI contents and fecal samples. *Indicates significant differences (p<0.05) between groups, **(p-0.001), ***(p<0.0001) and ****(p<0.00001).

#### Characterization of the small intestinal content by culturomics

The SI microbiota has not yet been the subject of culturomic study, a culture approach that involves the cultivation of as many bacterial species as possible through the use of enrichment steps, specific media, and incubation periods under both aerobic and anaerobic conditions. The culture-based analysis of the SI microbiota has the potential to facilitate the discovery of new species and the extensive study of isolated human gut bacteria that could have important implications in health and disease. Two SI contents recovered from two different healthy subjects were analyzed by culturomics. For the SI content of the first healthy subject, 733 colonies were isolated (666 in anaerobic and 67 in aerobic culture conditions). Subsequent identification of the isolates was facilitated by MALDI-TOF, resulting in precise species designation for 89% of the isolated strains. The remaining 11% of the isolates were identified through the sequencing of the entire 16S rRNA gene. The collective analysis yielded the identification of 70 distinct bacterial species (see Figure 7a and refer to the data in Supplementary Table 6). The majority of these isolates were classified into the phyla *Bacillota* and *Bacteroidota*, accounting for 60% and 21% of the identified species, respectively (refer to Figure 7b for further details). A total of four potential novel bacterial species belonging to the genera *Blautia, Extibacter, Flintibacter* and *Megaspharea* were identified based on their full-length 16S rRNA gene sequences. Their nucleotide similarities were less than 98.65% compared to publicly available and annotated 16S rRNA gene databases (EZbiocloud database update 2023.08.23) (see Figure 7d). The evaluation of the number of isolates and the final number of potentially newly identified species revealed the presence of one species for every 10.4 isolations. A second SI content was obtained from a different healthy subject and analyzed by culturomics. Overall, 736 colonies were isolated (716 in anaerobic and 20 in aerobic culture conditions). A total of 87% of the isolates could be identified by MALDI-TOF, leading to the identification of 40 bacterial species (Figure 7a and Supplementary Table 6). The majority of the 40 isolated bacterial species belonged to the phylum *Bacillota* and *Bacteroidota* (48% and 25% respectively; Figure 7c). For the SI content of this second healthy individual, we identified one potential new bacterial species belonging to the genus *Traorella* (Figure 7d). Combining both samples yielded a total of 90 unique bacterial species isolated and preserved during the course of the experiments (Figure 7d). Ten bacterial species were found in both samples (Figure 7e). Detailed growth conditions for each species are described in Supplementary Table 7. A comparison of the bacterial genera identified during culturomics with those detected by 16S metabarcoding revealed that 27 genera were exclusively detected by culturomics in sample 1 and 11 genera were identified in sample 2 (Supplementary Table 8). Conversely, 40 and 39 genera were exclusively identified through 16S metabarcoding in sample 1 and 2 respectively.

**Figure 7:**
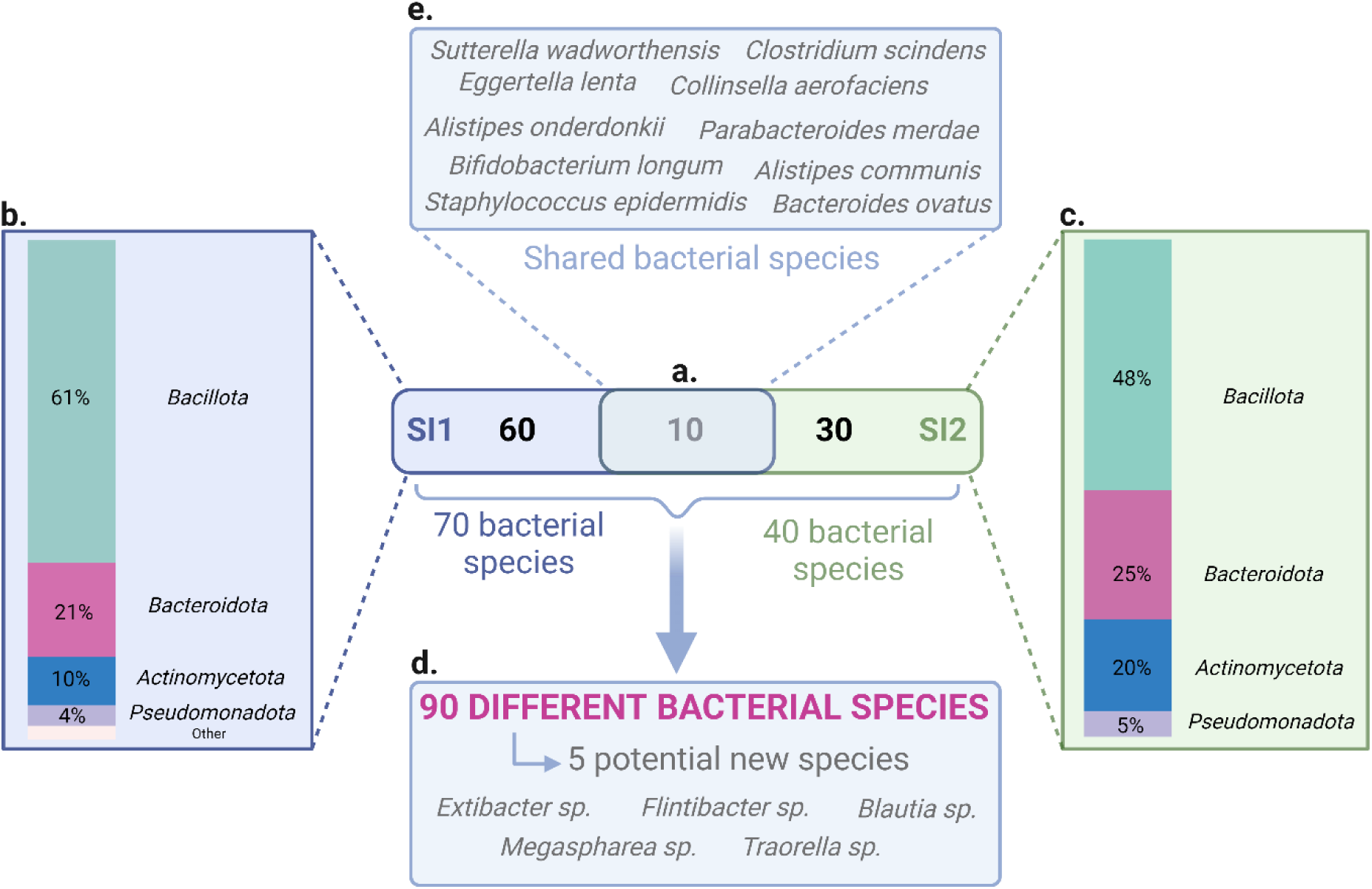
Isolated bacterial species by means of culturomics for small intestinal contents sampled from two different healthy volunteers. (a) Total number of microbial species identified for each sample. SI1: SI content 1 and SI2: SI content 2. (b) Percentage of identified species for each bacterial phylum for the SI content 1. “Other” contains two phyla: *Thermodesulfobacteriota* (1,4%) and *Campylobacterota* (1,4%). (c) Percentage of identified species for each bacterial phylum for the SI content 2. (d) Total number of distinct microbial species identified during both culturomics and including the number of the discovered new bacterial species. (e) Core bacterial species identified in both SI contents (figure created in BioRender).

## DISCUSSION

The SI is a key section of the GIT where essential process such as digestion and nutrient absorption occur. Unlike the fecal microbiome, which has been extensively studied and better characterized, the SI microbiome remains largely underexplored due to the difficulty of accessing sample. Indeed, the main challenge for studying the SI microbiome is its accessibility, usually requiring invasive methods to obtain SI contents samples. Here, we investigated the microbial and metabolite composition of SI contents sampled with a new sampling MD compared to corresponding fecal samples in 14 healthy subjects. We aimed to show that the new MD could significantly advance the characterization of the SI microbiome of healthy volunteers and that its future application in SI microbiome studies and the design of clinical trials will aid to eventually identify new diagnostic biomarkers of disease associated with the SI microbiome.

Our results demonstrate that the microbial composition of the SI differed significantly from that of the fecal samples, with a lower ASVs richness and α-diversity in the SI. This observation is consistent with previous studies highlighting that the SI microbiota is less diverse due to more restrictive physiological conditions (pH, flow rate, bile presence). The SI microbiota was generally predominated by Gram-positive genera such as *Schaalia*, *Gemella*, *Granulicatella* (Supplementary Table 3). Previous studies on the SI microbiota in healthy volunteers reported an overrepresentation of *Streptococcus*, *Veillonella*, *Gemella* and *Prevotella*^27,31,32^. Here, we also detected a trend that the relative abundance of *Streptococcus*, *Veillonella* and *Prevotella* was greater in SI contents compared to feces. It is imperative to note that a significant degree of inter-individual variation was detected within the SI microbiota group. Higher abundance of *Bifidobacterium* and *Faecalibacterium* was observed in some subjects of the SI content group compared to other subjects in our study (Figure 3d) and to the others clinical studies describing the SI microbiota^27,29,31,32^. The observed variations could be attributed to disparities in the designated sites of collection. Indeed, the MD is pH-sensitive and disintegrates at enteric pH^41^. Possible inter-individual pH variations^63^, buffer capacity^64–66^, intestinal fluid quantities and repartitions^67^ could influence the location where the MD disintegrates and therefore the SI sample collection site. Furthermore, it is plausible that the bacterial composition within the sampling module undergoes alterations during transit, as the MD is devoid of preservatives that would stabilize the bacterial composition. Consequently, the continued proliferation of certain bacterial species during transit has the potential to distort the microbial composition of the collection site. Moreover, baseline inter-individual differences in microbiome composition may further contribute to the variability observed across samples, complicating the distinction between biological variation and sampling-related effects. Procházková *et al.*, have recently shown the impact of gut physiology and the environment on the composition and metabolism of the gut microbiome, with the pH and transit time of each volunteer affecting the microbiome composition^68^.

To achieve an unprecedented characterisation of SI metabolome in healthy volunteers, we applied a combined approach integrating untargeted and semi-targeted metabolomics^47^. This strategy allowed us to determine the global metabolic profiles of SI contents and fecal samples, highlighting a distinct metabolic signature specific to each sample type. We identified key metabolites that were unique or significantly more abundant in one sample type compared to other, including TCA, CGA, and CA. Notably, BAs emerged as major discriminant metabolites identified through untargeted metabolomics, explaining many of the differences between the sample types. Using the semi-targeted metabolomics, we quantified 37 key metabolites and found that 15 of them were significantly more abundant in SI contents, with a large proportion corresponding to amino acids and BAs. The high quantity of BAs and amino acids observed in the SI contents likely reflect the intestine’s primary functions, such as digestion and nutrient absorption^27,38^. This finding aligns with previous reports by Folz *et al*., who also described elevated levels of BAs and protein breakdown products in the SI^38^. Amino acids, which are derived from the degradation of proteins and peptides, are absorbed predominantly in the SI, while primary BAs, secreted by the host in the duodenum, play a major role in emulsifying lipids and facilitating their absorption^69^. These functional roles explain their relatively high abundance in the SI microbiota.

Consistent with our findings, we showed that microbe-derived BAs were significantly more abundant in fecal samples, while host-derived BAs were predominantly concentrated in SI contents. Additionally, host/microbe-derived BAs appeared to be more abundant in SI contents, although this difference was only marginally significant (p=0.058), possibly due to intra-individual variations and the relatively small sample size. Notably, no significant difference was detected in the total BA concentration, likely because the subjects ingested the MD after a 10-hour fast. The predominance of host-derived BAs (such as GCA, TCA) in SI contents is expected since BAs are secreted in the duodenum via bile following meals^69^. Conversely, the higher abundance of microbe-derived BAs in feces likely reflects the increased microbial load and diversity in the colon, which promotes the transformation of primary BAs into secondary BAs by microbial enzymatic activity. BA concentrations can vary significantly due to several factors including dietary habits and circadian rhythm. For example, taurine- and glycine-conjugated BAs typically peak around mealtimes, while unconjugated BAs (CA, CDCA, LCA, DCA) peak around midnight and remain stable throughout the night^70^. Additionally, age-related shifts in the BA pool have been reported, with older individuals showing higher level of conjugated BAs, particularly taurine conjugates^71^. These variations make BA profiling across the healthy population challenging and highlights the need for clinical trials using a non-invasive MD to establish a robust and comprehensive characterization of the healthy SI metabolome.

Our study also highlights that the bacterial enzyme bile salt hydrolase (BSH) is capable of deconjugating not only taurine and glycine conjugates but also other amino acid conjugates. Importantly, most BSH- expressing bacteria are predominantly present in the large intestine. When examining the distribution of the predicted candidate BAs, we found differential abundances between SI and fecal samples. BAs function as key signaling molecules and microbiome metabolites that influence host-microbe interactions, and dysregulation of BA metabolism has been associated with various diseases, such as inflammatory bowel disease (IBD). In IBD, for example, alterations in BA metabolism can result in increased levels of primary BAs and decreased level of secondary BAs^72^, which may contribute to the dysregulation of BA receptors like Farnesoid X receptor (FXR). FXR plays crucial roles in regulating inflammation, autophagy, and immune responses^73^. Furthermore, the microbial transformation of BAs and their interaction with receptors have been linked to a wide range of metabolic, infectious, and neoplastic diseases^74,75^. Understanding these interactions could open new avenues for developing therapeutic strategies targeting BAs and their receptors in gastrointestinal and systemic diseases^76^.

To thoroughly characterize human SI microbial functions, a culturomics approach is useful. In this study, fast culturomics enabled the isolation and preservation of a diverse range of human SI gut bacteria, yielding a total of 90 species from two healthy subjects. Among these, we identified primarily anaerobic species, including several potentially novel microbial species (*Blautia sp.*, *Extibacter sp.*, *Flintibacter sp*., *Megasphaera sp.*, and *Traorella sp.*). By comparing the bacterial genera isolated via culturomics with those detected through 16S rRNA gene sequencing, we observed that some genera were uniquely identified by one method but not the other. In 2025, Diop *et al.*, also described the importance of combining culturomics with metagenomics, as 94 individual species were uniquely detected by culturomics^77^. This highlights the complementary nature of these approaches, as each method carries inherent biases. While sequencing techniques often underrepresent low-abundance microorganisms due to methodological factors such as DNA extraction and primer specificity^78^, culture techniques do not allow the isolation of all microbes^79^. Integrating culturomics with sequencing provide a more comprehensive view of the microbiota. One major advantage of culturomics lies in its ability to characterize isolates individually or in co-culture, enabling detailed phenotypic and genotypic characterization. This is particularly valuable for assessing the potential probiotic properties of specific bacterial strains.

Using a novel MD, we successfully performed an in-depth characterization of the small intestinal microbiome in healthy volunteers. By combining 16S rRNA gene sequencing, untargeted and semi-targeted metabolomics, BA-targeted metabolomics, and culturomics, our study demonstrates the feasibility and effectiveness of multi-omics analysis using samples collected with this novel sampling MD.

The microbial profiles and metabolomes exhibited marked differences between SI contents and fecal samples, particularly in terms of BA composition. SI content showed higher concentrations of host-derived and host/microbe-derived BAs compared to feces. This finding underscores the unique metabolic environment of the SI and highlights the potential of the MD as a groundbreaking tool for studying the SI microbiome in clinical trials. By enabling multi-omics analyses on non-invasively collected samples, this device could transform diagnostic approaches and biomarker discovery. Notably, even within a small cohort of healthy subjects, we identified specific SI biomarkers, such as host-derived BAs, emphasizing the clinical relevance of this innovative sampling method.

## Data Availability

Raw data for dataset are not publicly available to preserve individuals privacy under the European General Data Protection Regulation. Data supporting the findings of this study are available from the author A.L.G and T.S upon request.

## ACKNOWLEDGMENTS

We warmly thank Pr Max Maurin, Dr Aurélie Hennebique and Dr Camille Brunet from the Bacteriology Laboratory, Institute of Biology and Pathology, Grenoble Alpes University Hospital, for granting access to the MALDI-TOF mass spectrometer and for their valuable advice.

## ETHICAL STATEMENTS

This research was approved by the CHUGA institutional review board and authorized after its filing with the CNIL according to the French procedure for a monocentric study and has been granted ethical approval by the Personal Protection Committee (23 February 2022 and 9 March 2023) and, by the French National Agency for the Safety of Medicines and Health Products (ANSM) (2 June 2022 and 20 March 2023), and it has formally been registered as a study (NCT05477069).

## DISCLOSURE STATEMENT

A.T and T.S, as employee and CEO/co-founders of Pelican Health respectively, which markets intestinal sampling capsules, may face a conflict of interest due to their roles within the company. D.M, J.P.A and P.C are co-founder of Pelican Health. A.L.G is a co-founder for ALPIONER Therapeutics. P.C.D is an advisor and holds equity in Cybele, BileOmix and Sirenas and a Scientific co-founder, advisor and holds equity to Ometa, Enveda, and Arome with prior approval by UC-San Diego. P.C.D also consulted for DSM animal health in 2023. S.C.P, M.L and G.R are employees of Danone (France and Netherlands). All the other authors, declare that they have no conflict of interest.

## FUNDING

The study is supported by (1) the Direction de la Recherche Clinique et de l’Innovation (DRCI), CHUGA, Grenoble, France; (2) Association Nationale Recherche Technologie (ANRT) with a CIFRE fellowship n° 2021/0931, Paris, France; and (3) Pelican Health, Grenoble, France. (4) BPIFrance: i-Lab DOS0172081/00. We also acknowledge funding from NIDDK 1R01DK136117-01 granted to P.C.D.

## AUTHOR CONTRIBUTION

Author contributions were as follows: Conceptualization, A.T, P.C, J.P.A and T.S; Funding acquisition, T.S, A.S.S, P.C and D.M; Methodology, A.T, A.L.G and T.S; Performed research, A.T, C.P, S.B and A.C; Analyzed data, A.T, M.R.M, E.B., S.C.P and I.M ; Supervision, A.L.G, E.B and T.S; Writing original draft, A.T, E.B and T.S. All authors read, reviewed and approved the manuscript.

## DATA AVAILABILITY STATEMENT

Raw data for dataset are not publicly available to preserve individuals’ privacy under the European General Data Protection Regulation. Data supporting the findings of this study are available from the author A.L.G and T.S upon request.

## Notes

### Clinical Trial

NCT05477069

### Author Declarations

This research was approved by the Grenoble Alpes University Hospital institutional review board and authorized after its filing with the CNIL (Commission nationale de l'informatique et des libertes) according to the French procedure for a monocentric study and has been granted ethical approval by the Personal Protection Committee (CPP) (23 February 2022 and 9 March 2023) and, by the French National Agency for the Safety of Medicines and Health Products (ANSM) (2 June 2022 and 20 March 2023), and it has formally been registered as a study (NCT05477069).

