## Supplemental Figures 1 to 3 for "Profiling the human luminal small intestinal microbiome using a novel ingestible medical device"

### Supplementary Figures:

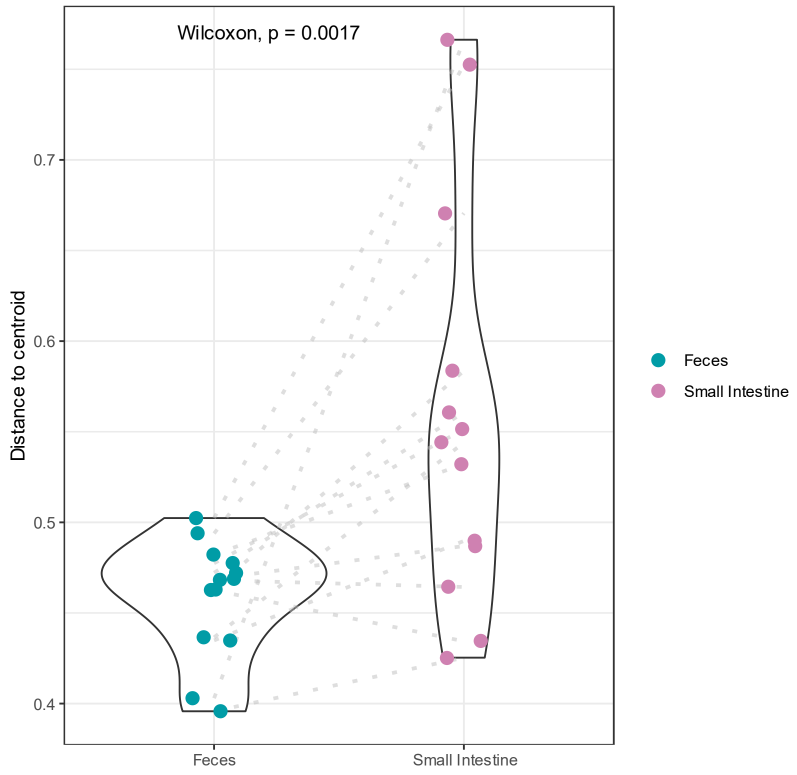


Supplementary figure 1: Permutational Analysis of Multivariate Dispersions (distance to the centroid) of small intestinal contents and feces using the PERMDISP function (R; vegan package). Differences in beta diversity among different areas or groups of samples were tested and results show a significant difference between the microbiota of the feces and SI samples.

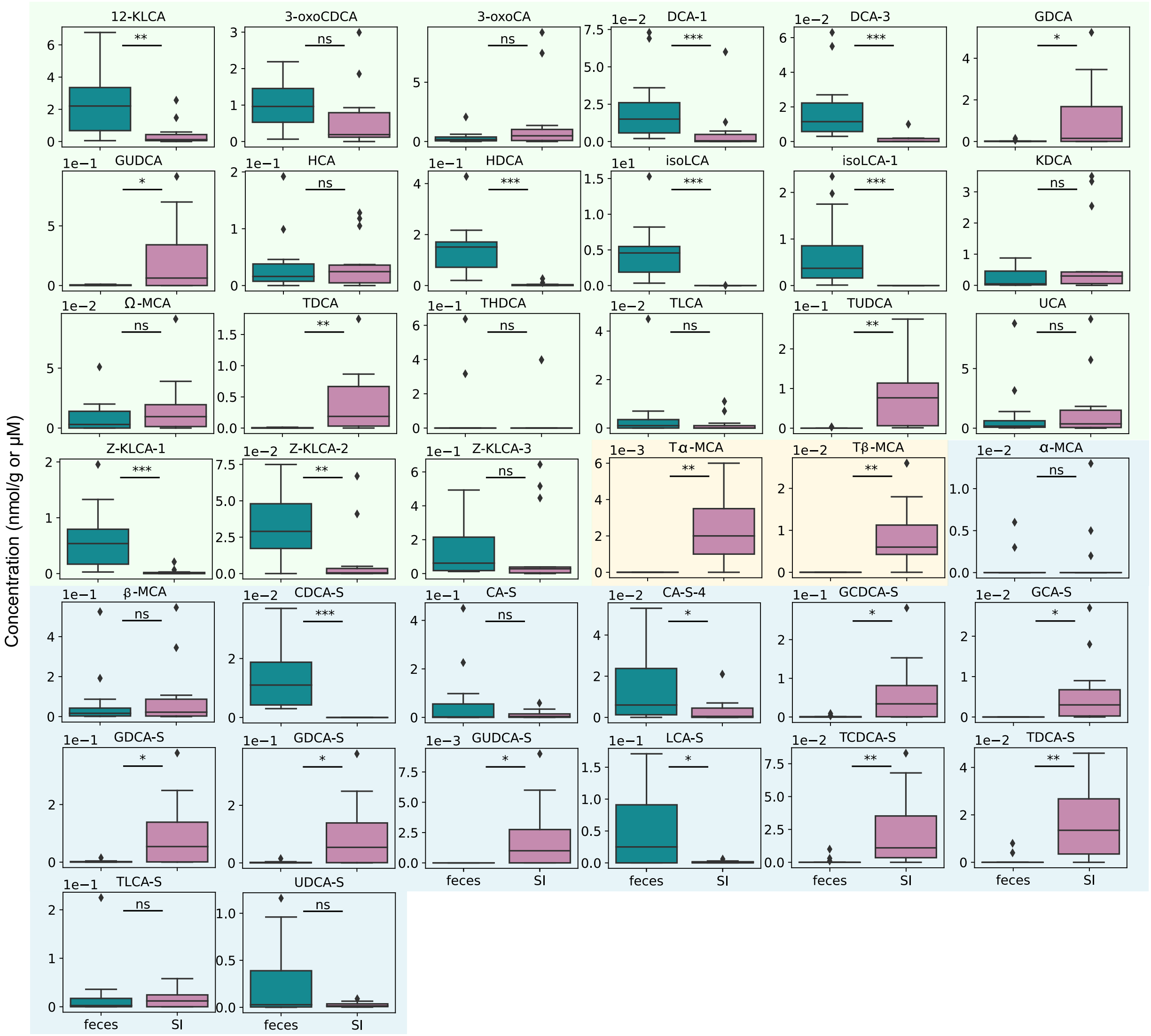


Supplementary figure 2: Boxplots of the quantified BAs in SI content versus feces. Green: microbe-derived BAs, yellow: host-derived BAs and blue: host/microbe-derived BAs. 12-KLCA: 12-Ketolithocholic acid; 3-oxoCDCA: 3-Oxochenodeoxycholic acid; 3-oxoCA: 3-Oxocholic acid; DCA-1: Deoxycholic acid like 1; DCA-3: Deoxycholic acid like 3; GDCA: Glycodeoxycholic acid; GUDCA: Glycoursodeoxycholic acid; HCA: Hyocholic acid; HDCA: Hyodeoxycholic acid; isoLCA: Isolithocholic acid; isoLCA-1: Isolithocholic acid like; KDCA: Ketodeoxycholic acid like; Ω-MCA : omega-Muricholic acid; TDCA: Taurodeoxycholic acid; THDCA: Taurohyodeoxycholic acid; TLCA: Taurolithocholic acid; TUDCA: Tauroursodeoxycholic acid; UCA: Ursocholic acid; Z-KLCA-1: Z-Ketolithocholic acid-like 1; Z-KLCA-2: Z-Ketolithocholic acid-like 2; Z-KLCA-3: Z-Ketolithocholic acid-like 3; GCDCA: Glycochenodeoxycholic acid; GCA: Glycocholic acid; Tα-MCA: Tauro-alpha-Muricholic acid; Tβ-MCA: Tauro-beta-Muricholic acid; α-MCA: alpha-Muricholic acid; β-MCA: beta-Muricholic acid; CDCA-S: Chenodeoxycholic acid-sulfate like; CA-S: Cholic acid-sulfate like; CA-S-4: Cholic acid sulfate like 4; CDCA-S: Glycochenodeoxycholic acid-sulfate like; GCA-S: Glycocholic acid-sulfate-like; GDCA-S: Glycodeoxycholic acid-sulfate like; GLCA-S-3: Glycolithocholic acid-sulfate like 3; GUDCA-S: Glycoursodeoxycholic acid-sulfate like; LCA-S: Lithocholic acid-sulfate-like; TCDCA-S: Taurochenodeoxycholic acid-sulfate like; TDCA-S: Taurodeoxycholic acid-sulfate like; TLCA-S: Taurolithocholic acid sulfate and UDCA-S: Ursodeoxycholic acid-sulfate like. Taurocholic acid- sulfate like and glycolithocholic acid have not been represented because their concentrations were below the limit of detection in both sample types. *Indicates significant differences ($p<0.05$) between groups, **($p<0.001$), ***($p<0.0001$) and ****($p<0.00001$).

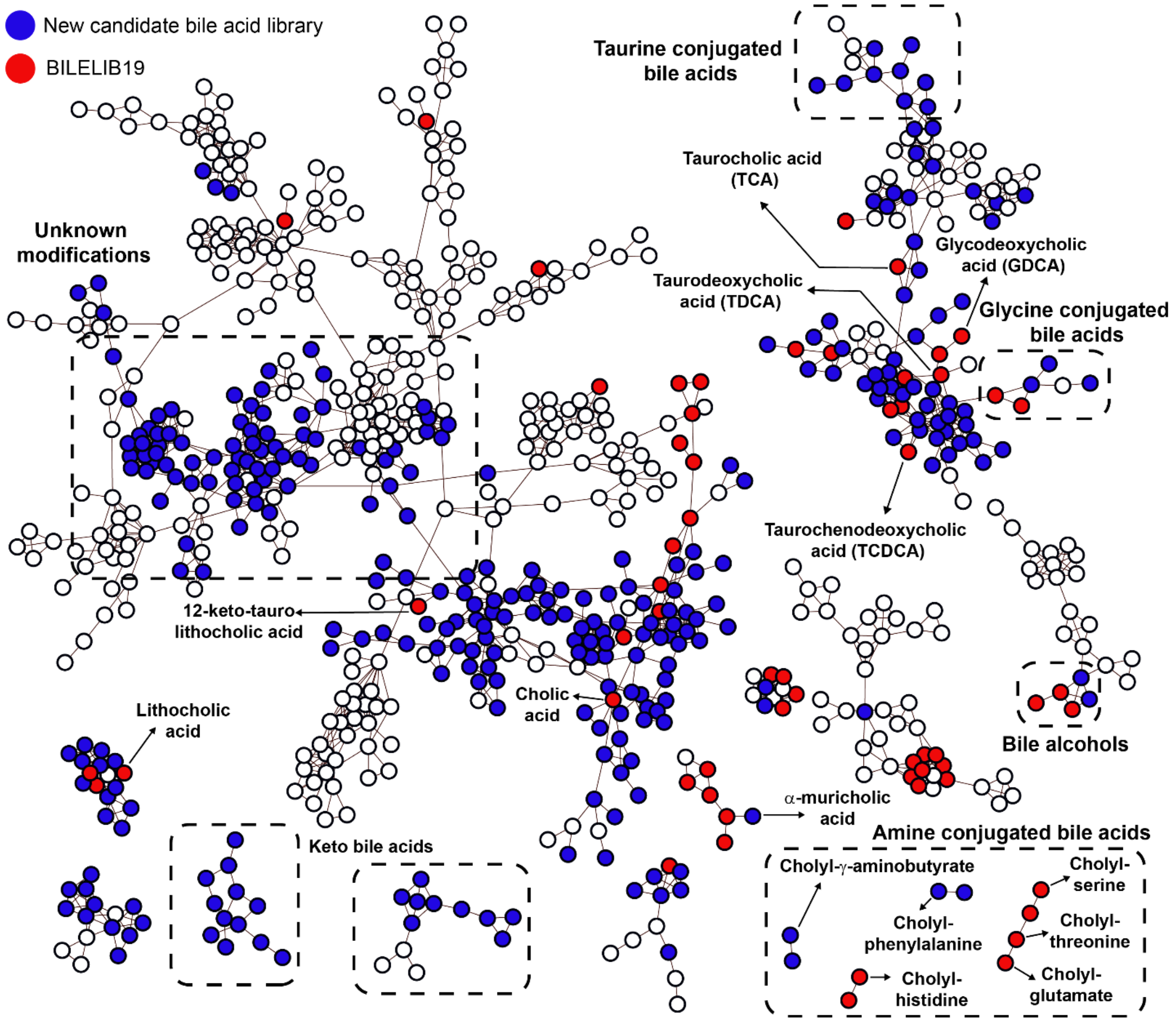


Supplementary figure 3: Feature-based molecular networking of annotated and non-annotated bile aids in fecal and SI samples of 14 patients. Molecular clusters containing spectral matches to two bile acid-specific MS2 libraries – the recently curated candidate library (blue nodes; 21,549 spectra are included in the library) and the BILELIB19 (red nodes; in total 5008 spectra are included in the library).
